# Technology And Its Role In Supporting Tuberculosis Treatment Adherence: A *Literature Review*

**DOI:** 10.1101/2021.12.22.21268290

**Authors:** Era Dorihi Kale, Moses Glorino Rumambo Pandin

## Abstract

Compliance with TB treatment has now become a problem that must be handled seriously because the high non-adherence rate will give a bad contribution to the success of TB treatment, including MDR-TB and also morbidity and mortality. Many innovations have been made to improve TB treatment adherence, one of which is using mobile-based technology. This article aims to explore the effectiveness of the technology used to improve treatment adherence in TB patients: types, ways of working, advantages, and limitations of each application. This is a systematic review through searching 3 databases, namely Scopus, WoS, and Science Direct. Some of the advantages in applying technology to improve TB treatment adherence are easy to use if you understand how to operate tools/applications are cost-effective because they reduce transportation costs in reaching remote areas or in conditions of transportation difficulties such as after a disaster, the use of this technology provides patient satisfaction in treatment and facilitates the involvement of the family/support system in the treatment of patients. Several things must be considered (limitations) of the technology to be used, including experts, patient knowledge and skills, economic condition, electricity availability, and whether the technology used will not increase the burden on patients related to the stigma of TB disease. We can conclude that the use of technology is indeed very good in supporting the improvement of TB treatment adherence, but the selection of this application must pay attention to the characteristics of the population as well as the advantages and limitations of each application.

## INTRODUCTION

Tuberculosis (TB) is an infectious disease that causes the most deaths with a single infectious cause, can affect anyone. Until now, a quarter of the world’s population has been infected with this disease, of which 44% came from Southeast Asia (WHO, 2020). Globally, the target of achieving TB treatment has not been achieved yet (WHO, 2020)(Byonanebye *et al*., 2021), with a high rate of non-adherence to treatment in TB patients, will greatly affect the increase in the number of MDR-TB (Multi Drugs Resistant). The factors that influence the level of adherence of TB patients in treatment are lack of knowledge, economic factors, stigma, lack of social support, drug side effects, long duration of treatment. (Gebreweld *et al*., 2018), disbelief that medicine can heal (Azizi, Karimy, and Salahshour, 2018), difficult access to treatment facilities (Ruru *et al*., 2018) and lack of family support as supervisors to help monitor patients taking medication (Chen *et al*., 2020).

Compliance with taking medication for TB patients can be improved through the presence of a drug-taking supervisor (DTS). DTS can come from family members, closest relatives, or health workers who are committed and have time to supervise patients during the treatment period, but DTS also have limitations, namely: busyness, boredom and the number of people willing to become PMOs is decreasing. To overcome this, technology has been developed that aims to improve patient compliance with taking drugs and to keep patients continuing treatment according to the established program. Most of these smartphone-based applications have been tested directly in the community with varying effectiveness (Wang *et al*., 2019)(Iribarren *et al*., 2021). The use of this technology is not only aimed at patients but also at health workers who serve TB patients (Patel *et al*., 2020). The applications developed are diverse and have their respective approaches so that each application has its advantages and disadvantages (Byonanebye *et al*., 2021). A lot of research has been done to measure the effectiveness of each application, therefore this article will discuss it as a whole.

## OBJECTIVE

Explore the effectiveness of the technology used to improve treatment adherence in TB patients: types, ways of working, advantages, and limitations of each application.

## METHOD

This article is a systematic literature review that focuses on the use of technology in improving treatment adherence in TB patients. The method used is Literature Review. The journals taken are international reputable journals with appropriate themes. The databases used in this literature search are Scopus, Web of Science, and Science Direct. In the article search, the author uses the keywords: Technology, Tuberculosis, Adherence. After conducting a literature search in 3 databases with the keywords above, the selected articles are published in the last 3 years (2019. 2020, and 2021) and full research articles (not literature reviews). The researcher will then make a selection based on the title, then based on the abstract, and finally the full-text selection. The results of the selection will be used as articles to be reviewed. The data were analyzed descriptively by the research objectives that have been set.

**Figure 1.**
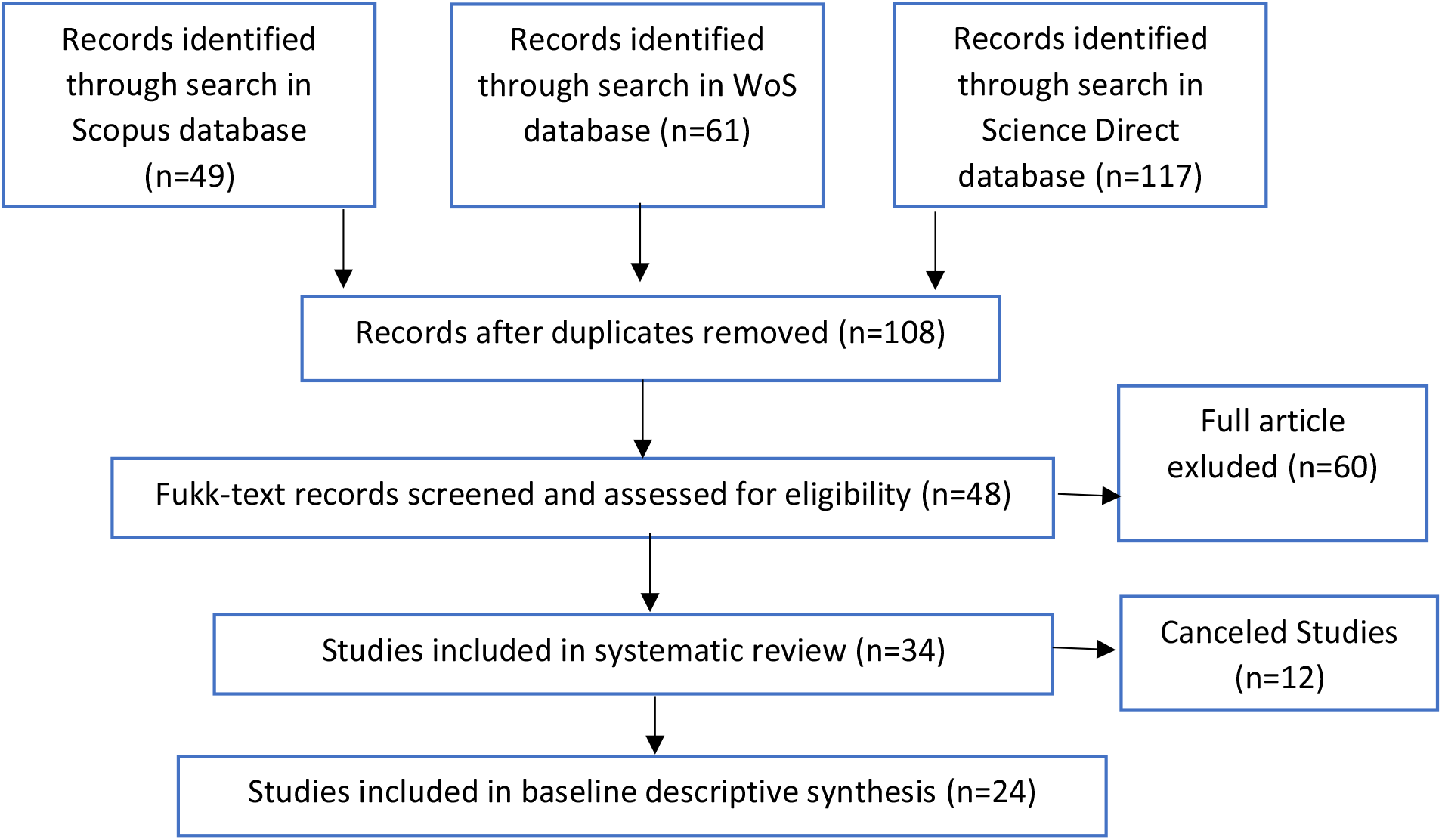
Article Selection Flow (PRISMA 2020)

## RESULT

The use of technology to monitor TB treatment adherence is not a very new thing, but many innovations have been developed to make this technology increasingly able to answer the need to improve TB patient treatment adherence. The table below is a summary of the applications that have been evaluated through research.

**Table 1.**
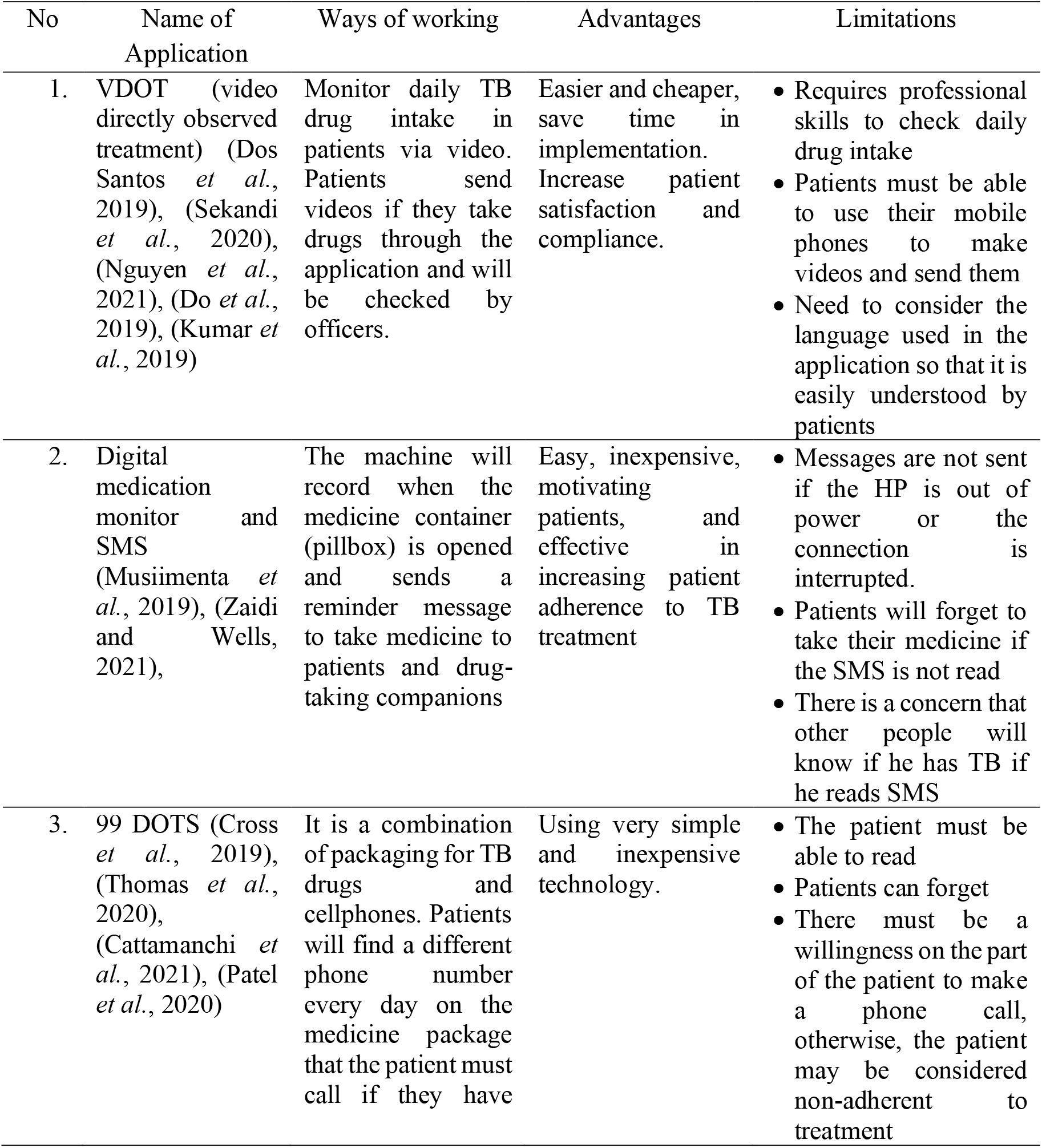
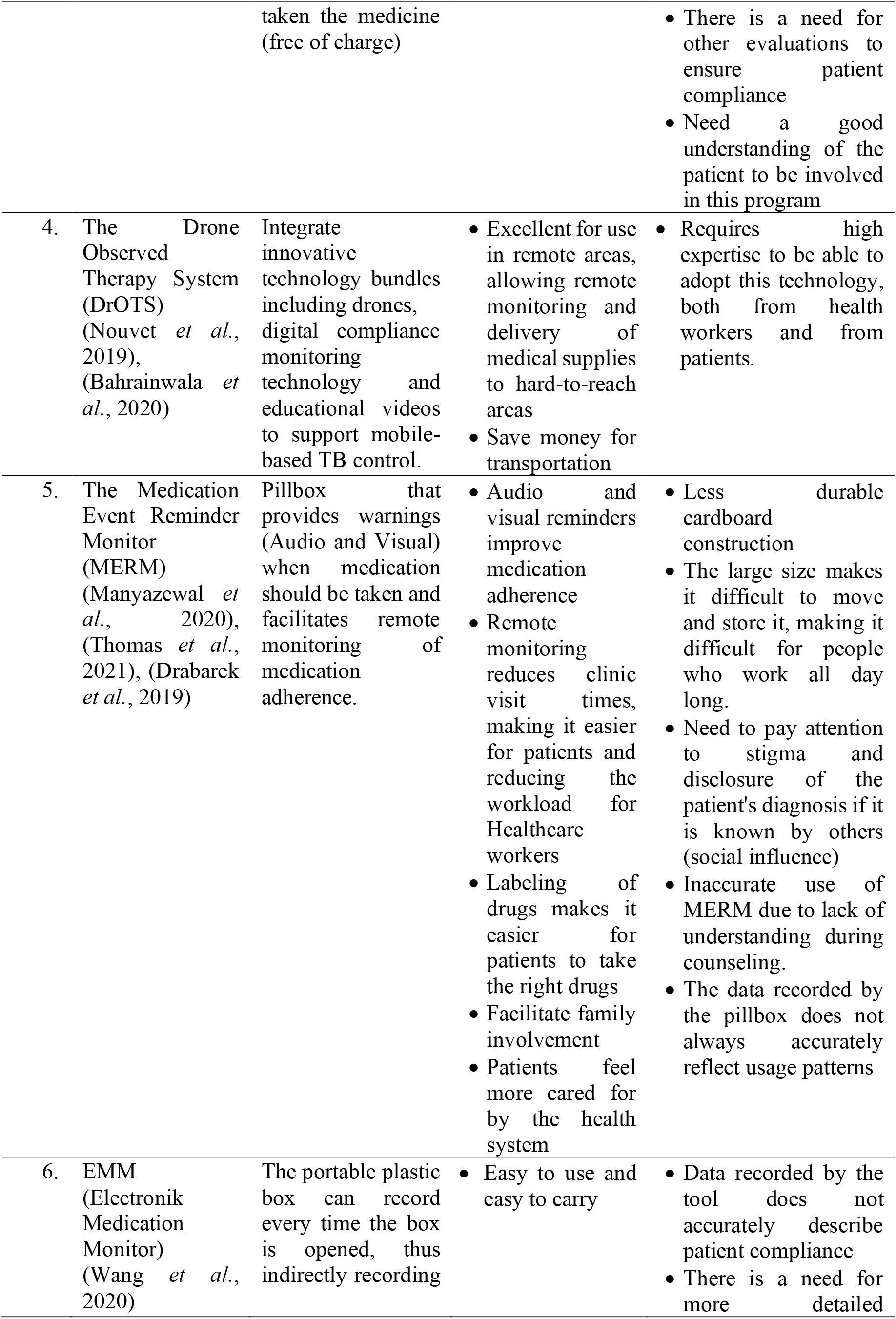
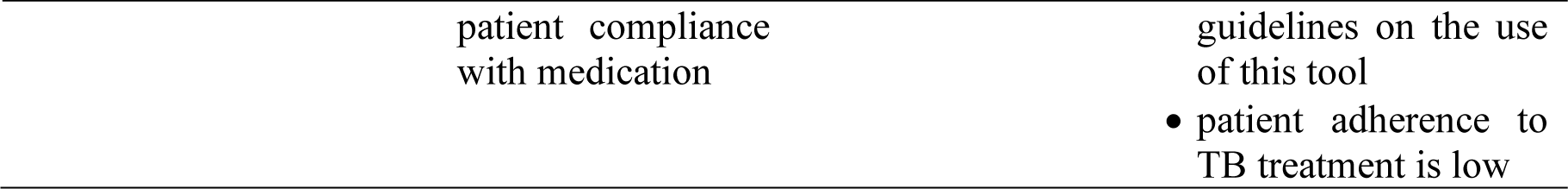
Application to improve TB treatment adherence (advantages and limitations)

## DISCUSSION

The use of mobile/smartphone-based technology has been shown to provide a significant increase in TB treatment adherence. Based on the results above, it can be seen that there are 6 types of methods to improve patient compliance, namely: VDOT (video directly observed treatment), Digital medication monitor and SMS, 99 DOTS, The Drone Observed Therapy System (DrOTS), The Medication Event Reminder Monitor (MERM) and EMM (Electronic Medication Monitor), although based on research results show that the effectiveness of each tool is different depending on population characteristics and also local socio-cultural, Some of the advantages in applying technology to improve TB treatment adherence are: easy use if you understand how to operate tools/applications, are cost-effective because they reduce transportation costs in reaching remote areas or in conditions of transportation difficulties such as after a disaster, the use of this technology provides patient satisfaction in treatment and facilitates the involvement of the family/support system in the treatment of patients.

Apart from the benefits, several things must be considered (limitations) of the technology to be used, including whether the tool/application used will require experts with special abilities, the patient’s ability to run the application in terms of knowledge, skills, and economically (whether the patient can afford to buy a compatible cellphone/smartphone for the application. Another factor that must be considered is how to respond to the condition if the patient runs out of power/damage to the cellphone, for example by automatically diverting messages to the HP support system (people closest to you)), so that the reminder to take medication will not be interrupted, it is also added how to prepare patients and the support system to really understand the proper use of the application, and the need for a detailed manual on the application manual. menu research suggests that the patient may forget his or her treatment plan if there is no reminder. The last thing to consider is whether the application used will exacerbate TB stigma in the community, related to disclosure of TB status due to reminders that are known by others.

## CONCLUSION

The use of technology is indeed very good in supporting the improvement of TB treatment adherence, but the selection of this application must pay attention to the characteristics of the population as well as the advantages and limitations of each application.

## Data Availability

All data produced in the present work are contained in the manuscript

## REFERENCES

1. Azizi, N., Karimy, M. and Salahshour, V. N. (2018) ‘Determinants of adherence to tuberculosis treatment in Iranian patients: Application of health belief model’, Journal of Infection in Developing Countries, 12(9), pp. 706–711. doi: 10.3855/jidc.9653.

2. Bahrainwala, L. et al. (2020) ‘Drones and digital adherence monitoring for community-based tuberculosis control in remote Madagascar: A cost-effectiveness analysis’, PLoS ONE, 15(7 July), pp. 1–19. doi: 10.1371/journal.pone.0235572.

3. Bommakanti, K. K. et al. (2020) ‘Requiring smartphone ownership for mHealth interventions: Who could be left out?’, BMC Public Health, 20(1), pp. 1–9. doi: 10.1186/s12889-019-7892-9.

4. Byonanebye, D. M. et al. (2021) ‘Impact of a mobile phone-based interactive voice response software on tuberculosis treatment outcomes in Uganda (CFL-TB): a protocol for a randomized controlled trial’, Trials, 22(1), pp. 1–13. doi: 10.1186/s13063-021-05352-z.

5. Cattamanchi, A. et al. (2021) ‘Digital adherence technology for tuberculosis treatment supervision: A stepped-wedge cluster-randomized trial in Uganda’, PLoS Medicine, 18(5), pp. 1–15. doi: 10.1371/journal.pmed.1003628.

6. Chen, X. et al. (2020) ‘The effects of family, society and national policy support on treatment adherence among newly diagnosed tuberculosis patients: a cross-sectional study’, pp. 1–11. doi: 10.21203/rs.3.rs-25273/v1.

7. Cross, A. et al. (2019) ‘99DOTS: A low-cost approach to monitoring and improving medication adherence’, ACM International Conference Proceeding Series. doi: 10.1145/3287098.3287102.

8. Do, D. et al. (2019) ‘Change in patient comfort using mobile phones following the use of an app to monitor tuberculosis treatment adherence: Longitudinal study’, JMIR mHealth and uHealth, 7(2), pp. 1–8. doi: 10.2196/11638.

9. Drabarek, D. et al. (2019) ‘Implementation of Medication Event Reminder Monitors among patients diagnosed with drug susceptible tuberculosis in rural Viet Nam: A qualitative study’, PLoS ONE, 14(7), pp. 1–11. doi: 10.1371/journal.pone.0219891.

10. Gebreweld, F. H. et al. (2018) ‘Factors influencing adherence to tuberculosis treatment in Asmara, Eritrea: A qualitative study’, Journal of Health, Population and Nutrition, 37(1), pp. 1–9. doi: 10.1186/s41043-017-0132-y.

11. Iribarren, S. et al. (2021) ‘Mobile tuberculosis treatment support tools to increase treatment success in patients with tuberculosis in argentina: Protocol for a randomized controlled trial’, JMIR Research Protocols, 10(6). doi: 10.2196/28094.

12. Kumar, A. A. et al. (2019) ‘Mobile health for tuberculosis management in South India: Is video-based directly observed treatment an acceptable alternative?’, JMIR mHealth and uHealth, 7(4). doi: 10.2196/11687.

13. Manyazewal, T. et al. (2020) ‘Electronic pillbox-enabled self-administered therapy versus standard directly observed therapy for tuberculosis medication adherence and treatment outcomes in Ethiopia (SELFTB): Protocol for a multicenter randomized controlled trial’, Trials, 21(1), pp. 1–13. doi: 10.1186/s13063-020-04324-z.

14. Musiimenta, A. et al. (2019) ‘Digital monitoring technologies could enhance tuberculosis medication adherence in Uganda: Mixed methods study’, Journal of Clinical Tuberculosis and Other Mycobacterial Diseases, 17, p. 100119. doi: 10.1016/j.jctube.2019.100119.

15. Nguyen, L. H. et al. (2021) ‘Assessing private provider perceptions and the acceptability of video observed treatment technology for tuberculosis treatment adherence in three cities across Viet Nam’, PLoS ONE, 16(5 May), pp. 1–15. doi: 10.1371/journal.pone.0250644.

16. Nouvet, E. et al. (2019) ‘Perceptions of drones, digital adherence monitoring technologies and educational videos for tuberculosis control in remote Madagascar: A mixed-method study protocol’, BMJ Open, 9(5). doi: 10.1136/bmjopen-2018-028073.

17. Patel, D. et al. (2020) ‘Iterative adaptation of a tuberculosis digital medication adherence technology to meet user needs: Qualitative study of patients and health care providers using human-centered design methods’, JMIR Formative Research, 4(12), pp. 1–14. doi: 10.2196/19270.

18. Ruru, Y. et al. (2018) ‘Factors associated with non-adherence during tuberculosis treatment among patients treated with DOTS strategy in Jayapura, Papua Province, Indonesia’, Global Health Action, 11(1). doi: 10.1080/16549716.2018.1510592.

19. Dos Santos, L. R. A. et al. (2019) ‘The perception of health providers about an artificial intelligence applied to Tuberculosis video-based treatment in Brazil: A protocol proposal’, Procedia Computer Science, 164, pp. 595–601. doi: 10.1016/j.procs.2019.12.225.

20. Sekandi, J. N. et al. (2020) ‘Video directly observed therapy for supporting and monitoring adherence to tuberculosis treatment in uganda: A pilot cohort study’, ERJ Open Research, 6(1). doi: 10.1183/23120541.00175-2019.

21. Thomas, B. E. et al. (2020) ‘Evaluation of the Accuracy of 99DOTS, a Novel Cellphone-based Strategy for Monitoring Adherence to Tuberculosis Medications: Comparison of DigitalAdherence Data with Urine Isoniazid Testing’, Clinical Infectious Diseases, 71(9), pp. pE513–E516. doi: 10.1093/cid/ciaa333.

22. Thomas, B. E. et al. (2021) ‘Acceptability of the medication event reminder monitor for promoting adherence to multidrug-resistant tuberculosis therapy in two indian cities: Qualitative study of patients and health care providers’, Journal of Medical Internet Research, 23(6), pp. 1–18. doi: 10.2196/23294.

23. Wang, N. et al. (2019) ‘Using electronic medication monitoring to guide differential management of tuberculosis patients at the community level in China’, BMC Infectious Diseases, 19(1), pp. 1–9. doi: 10.1186/s12879-019-4521-2.

24. Wang, N. et al. (2020) ‘Electronic medication monitor for people with tuberculosis: Implementation experience from thirty counties in China’, PLoS ONE, 15(4), pp. 1–14. doi: 10.1371/journal.pone.0232337.

25. WHO (2020) Global Tuberculosis Report 2020.

26. Zaidi, H. A. and Wells, C. D. (2021) ‘Digital health technologies and adherence to tuberculosis treatment’, Bulletin of the World Health Organization, 99(5), pp. p323-323A. doi: 10.2471/BLT.21.286021.

